# Longitudinal Brain Atrophy Patterns in Dementia and Cognitive Decline: the Framingham Heart Study

**DOI:** 10.64898/2026.04.28.26351854

**Authors:** Habbiburr Rehman, Qiushan Tao, Jackson Nolan, Nuzulul Kurniansyah, Ting Fang Alvin Ang, Paul K. Crane, Shubhabrata Mukherjee, Andrew J. Saykin, Emily H. Trittschuh, Thor D. Stein, Jesse Mez, Rhoda Au, Lindsay A. Farrer, Douglas N. Greve, Xiaoling Zhang, Wei Qiao Qiu

## Abstract

**Background:** Characterizing longitudinal patterns of brain atrophy that distinguish Alzheimer’s disease (AD) and related neurodegeneration along with normative aging remains a major challenge. We aimed to identify data-driven longitudinal brain atrophy components and evaluate their associations with plasma AD biomarkers and cognitive outcomes in a community-based cohort.

**Methods:** We analyzed 756 MRI scans from 300 participants in the Framingham Heart Study (mean 2.52 scans per participant; range 2–4). Linear mixed-effects models were used to identify MRI features associated with diagnostic group (cognitively normal [CN], mild cognitive impairment [MCI], and dementia). Significant features (n=211) were entered into a longitudinal multivariate decomposition framework (ANOVA Simultaneous Component Analysis with Assorted Linear functions; ALASCA) to derive principal components (PCs) capturing patterns of structural change over time. Associations between PCs and plasma AD biomarkers (p-Tau_181_, total Tau(t-Tau), glial fibrillary acidic protein [GFAP], neurofilament light chain [NfL], amyloid-β40 [Aβ_40_], and amyloid-β42 [Aβ_42_]) were evaluated using multivariable mixed-effects models adjusted for age, sex, education, and APOE ε4 status. Cognitive measures and neuroethological measures in a subset were used to assess the functional relevance and biological associations, respectively.

**Results:** The first three PCs explained ∼95% of the variance within the modeled MRI feature (n=211) set (PC1: 75.8%, PC2: 13.8%, PC3: 5.4%). PC1 captured medial temporal atrophy involving hippocampal subfields and basolateral amygdala and was associated with worse cognition and higher plasma AD biomarkers. Neuropathological analyses showed stronger associations of PC1-related atrophy with AD-related tau pathology in the absence of concomitant TDP-43 pathology. In contrast, PC2 reflected diffuse cortical gray–white matter contrast alterations across association cortices and showed distinct associations with biomarkers and cognition compared to PC1, consistent with overlapping aging- and neurodegeneration-related processes. PC3 showed limited variance and no consistent associations.

**Conclusion:** Longitudinal MRI-derived components capture distinct patterns of brain structural change associated with neurodegeneration. Medial temporal trajectories are closely associated with AD and related dementia, whereas cortical alterations likely reflect mixed aging- and disease-related processes. Integration of structural MRI with plasma biomarkers provides complementary information on disease expression and heterogeneity, supporting multimodal approaches for disease characterization and risk stratification.

## 1 Introduction

Alzheimer’s disease (AD) is the leading cause of dementia and a growing public health challenge in aging populations. Early detection is critical for effective intervention, yet diagnosis during the preclinical and mild cognitive impairment (MCI) stages remains difficult due to clinical heterogeneity and overlap with normal aging and comorbid conditions.^1–5^ Although cerebrospinal fluid (CSF) biomarkers and positron emission tomography (PET) imaging provide valuable insights into amyloid and tau pathology, their high-cost, invasiveness, and limited accessibility constrain widespread clinical use.^5–8^ Blood-based biomarkers have emerged as promising alternatives^9,10^, but their interpretation in terms of brain pathological changes remains unclear.

Structural magnetic resonance imaging (MRI) offers a non-invasive, widely available tool for assessing neurodegeneration, with hippocampal atrophy serving as one of the most robust imaging markers of AD.^5–8^ Notably, hippocampal atrophy detected by MRI, together with abnormal CSF biomarker profiles, has been demonstrated to predict long term cognitive decline and associated with increasing AD pathology. However, most prior MRI studies rely on cross-sectional measures or global volumetric summaries, which fail to capture the dynamic and heterogeneous nature of brain atrophy over time.

Emerging evidence indicates that AD progression involves spatially and temporally distinct patterns of brain change, particularly within limbic structures such as hippocampal subfields and the amygdala.^11,12^ These regions are critical for memory and emotional processing but remain underexplored in longitudinal and population-based studies.^13,14^ Moreover, AD frequently co-occurs with other pathologies^11,15^, including limbic-predominant age-related TDP-43 encephalopathy (LATE), which contributes to clinical and neuroanatomical heterogeneity.^16,17^ However, longitudinal brain changes in pure AD compared with mixed AD and LATE remain incompletely characterized.

While data-driven longitudinal methods can capture such trajectories, few studies have integrated these approaches with plasma biomarkers and cognition in community-based cohorts.^4,18^ To address these gaps, we used longitudinal MRI data from the Framingham Heart Study (FHS) to identify data-driven patterns of brain structural change and examined their associations with plasma biomarkers, cognitive outcomes, and neuropathology across the spectrum from normal aging to dementia.

## 2 Method

### 2.1 Study participants

Data was obtained from Generation 1 (Original) and Generation 2 (Offspring) participants of the FHS, a community-based prospective cohort in Framingham, Massachusetts. Participants with longitudinal structural MRI data (n=1,256) processed using FreeSurfer-V7.5 were considered. After applying exclusion criteria, the final analytic sample included 726 MRI scans from 300 participants (**Figure-1; sTable-1**). Each participant contributed at least two scans (mean=2.52,range=2–4). The last MRI scan was performed in June 2015; by that time, no participants had developed dementia, and only a few had MCI (**sFigure-1**). Participants were subsequently followed longitudinally through October 2024 for incident dementia diagnosis. Additional details on cohort selection are provided in the **sMethod**. All participants provided informed consent, and the study was approved by the Boston University Institutional Review Board.

**Figure 1:**
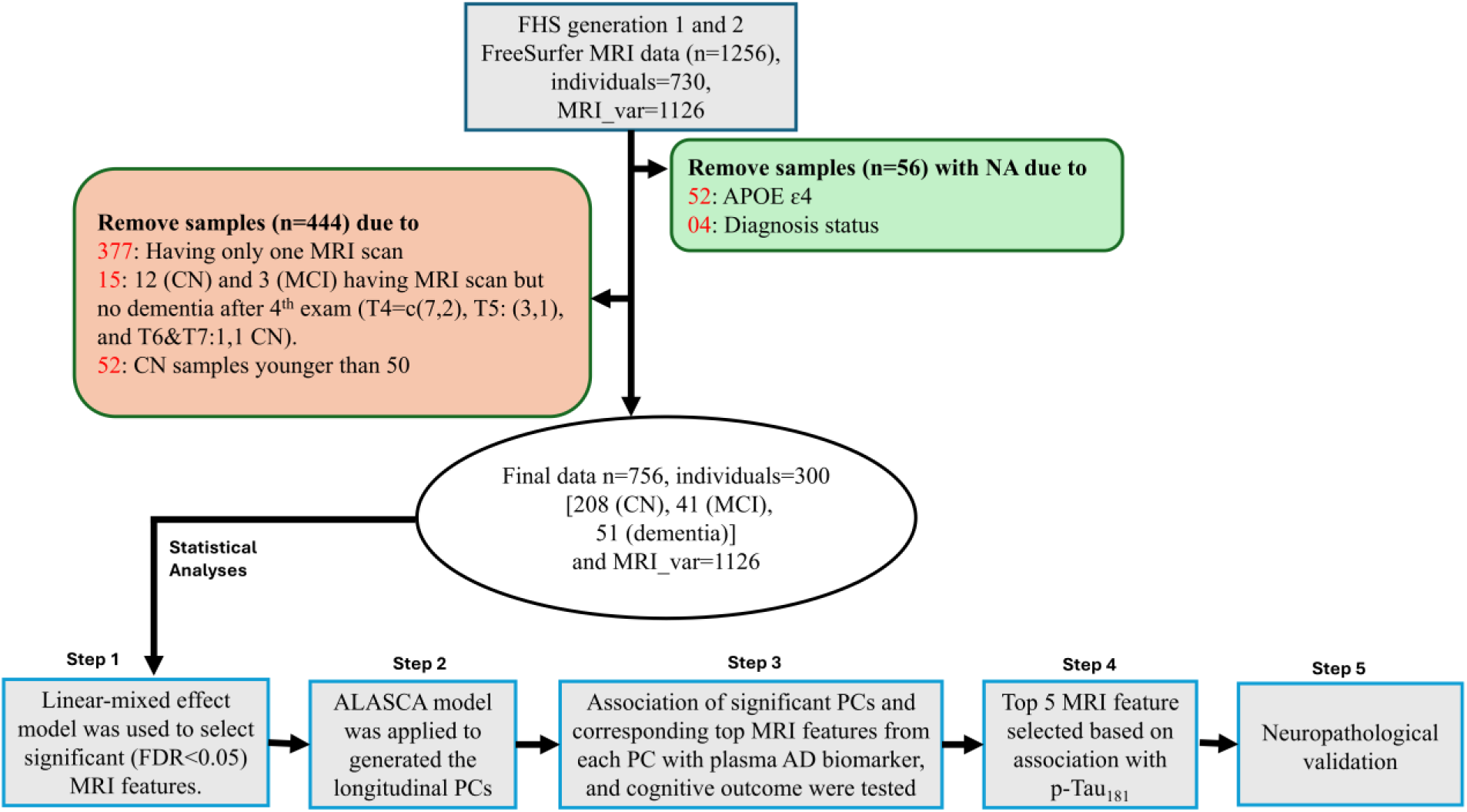
Schematic overview of the study design. Abbreviations: FHS= Framingham Heart Study; n=sample size; CN=Cognitive normal; MCI=Mild cognitive impairment; PCs=Principal components; MRI_var= MRI variables

### 2.2 Cognitive assessment

Cognitive status was assessed longitudinally in the FHS using standardized procedures. Dementia and MCI diagnoses were adjudicated by a multidisciplinary panel at least one neurologist and one neuropsychologist using established clinical criteria.^19–21^ Domain-specific cognitive factor scores (memory, language, executive function) were derived and harmonized across examination cycles.^22,23^ The most recent available cognitive assessment was used for association analyses to approximate cognitive status closest to the time of MCI or dementia onset. Additional details are provided in the **sMethod**.

### 2.3 MRI acquisition and processing

Structural T1-weighted MRI scans were processed using FreeSurfer-V7.5. Cortical thickness, surface area, regional volumes, hippocampal subfields, amygdalar nuclei, and gray–white matter contrast measures were extracted.^24–26^ Quality assurance included visual inspection and standardized rating procedures. Estimated total intracranial volume(eTIV) was derived and used for head-size normalization.^27^ Detail description available in the **sMethod**.

### 2.4 Plasma AD biomarkers

Plasma biomarkers were measured in the Offspring cohort using ultrasensitive single-molecule array (Simoa) assays and included phosphorylated tau 181 (p-Tau_181_), total tau (t-Tau), glial fibrillary acidic protein (GFAP), neurofilament light chain (NfL), amyloid-β40 (Aβ_40_), and amyloid-β42 (Aβ_42_). Biomarker values were log-transformed to achieve the normality.^28,29^ Detail description available in the **sMethod**.

### 2.5 Neuropathology

Neuropathology was assessed in a subset of FHS participants with postmortem brain tissue. Neurofibrillary tangle pathology was quantified using Braak staging. In addition, phosphorylated TDP-43 pathology and limbic age-related TDP-43 encephalopathy (LATE) were evaluated using established consensus criteria.^30,31^ Detailed methods are provided in the **sMethod**.

### 2.6 Statistical analyses

To identify MRI features showing disease-relevant differences across diagnostic groups, we fitted separate linear mixed-effects models for each feature. MRI measures were modeled as outcomes, with diagnostic group as the primary predictor and age at MRI, sex, education, and APOE ε4 status as covariates. A subject-specific random intercept accounted for repeated measurements. Features significant after FDR correction (FDR<0.05) were retained for multivariate trajectory modeling.

Longitudinal atrophy trajectories were derived using a multivariate decomposition framework, Assorted Linear functions for ANOVA–Simultaneous Component Analysis(ALASCA)^32^ This approach models time, group, and time-by-group interaction effects and decomposes them into principal components (PCs) representing shared longitudinal patterns of structural change (**sMethod**). Associations between ALASCA-derived PCs, plasma AD biomarkers, and cognitive outcomes were evaluated using multivariable linear mixed-effects models adjusted for age at MRI, sex, education, and APOE ε4 carrier status. Because clinical dementia diagnosis, which incorporates cognitive test performance, was included in the derivation of ALASCA-derived principal components, subsequent associations with cognitive outcomes were not fully independent and were interpreted as descriptive measures of the functional relevance of the identified imaging patterns. Detail statistical plan available in the **sMethod**.

## 3 Results

### 3.1 Participant characteristics

**Table-1** presents the baseline demographic and clinical characteristics of the study participants. The analytic sample included 300 individuals [CN=208,MCI=41, and dementia=51] (**Table-1**). Age at first MRI scan (p=7.06e-12) and sex (p=0.026) were significantly associated with diagnostic status. While APOE ɛ4 genotype tended to be associated with dementia education, whereas, education, and eTIV were not in this cohort (**Table-1**). In total, we analyzed 756 longitudinal MRI scans (with at least two and maximum four brain MRI scans) across all time points: CN=533, MCI=102, and dementia=121 (**sTable-1**). Most samples were from the Generation 2 cohort (n=657), with a smaller subset from the Generation 1 cohort (n=99) (**sTable-1**). This study included 38 individuals who developed AD dementia and 13 who developed other types of dementia (**Table-1**). All MRI scans were done on an average of approximately 5 and 8 years before the onset of MCI and dementia, respectively (**sTable-1**).

**Table 1:** Demographic and clinical characterstics of the study participants.

| Characteristic | Overall N = 300<sup>1</sup> | CN<br>N = 208<sup>1</sup> | MCI<br>N = 41<sup>1</sup> | Dementia<br>N = 51<sup>1</sup> | p-value<sup>2</sup> |
| --- | --- | --- | --- | --- | --- |
| **Age at first MRI** | 65.73 (9.46) | 63.23 (7.91) | 71.13 (10.90) | 71.59 (9.79) | 7.06e-12 |
| **Sex** |  |  |  |  | 0.026 |
| Male | 125 (41.67) | 97 (46.63) | 14 (34.15) | 14 (27.45) |  |
| Female | 175 (58.33) | 111 (53.37) | 27 (65.85) | 37 (72.55) |  |
| **Education** |  |  |  |  | 0.207 |
| High school graduate or not (0) | 99 (33.00) | 61 (29.33) | 19 (46.34) | 19 (37.25) |  |
| Some college (1) | 89 (29.67) | 62 (29.81) | 11 (26.83) | 16 (31.37) |  |
| College graduate (2) | 112 (37.33) | 85 (40.87) | 11 (26.83) | 16 (31.37) |  |
| **APOE ε4** |  |  |  |  | 0.102 |
| 0 | 251 (83.67) | 176 (84.62) | 37 (90.24) | 38 (74.51) |  |
| 1 | 49 (16.33) | 32 (15.38) | 4 (9.76) | 13 (25.49) |  |
| **eTIV (mm<sup>3</sup>)** | 1,565,523.94<br>(176,058.17) | 1,571,752.85<br>(180,936.76) | 1,567,002.15<br>(172,221.26) | 1,538,931.38<br>(158,778.22) | 0.491 |
| **Plasma AD Biomarkers** (Exam 08/09) |  |  |  |  |  |
| p-Tau<sub>181</sub> | 0.98 (0.47) | 0.91 (0.44) | 1.30 (0.52) | 1.31 (0.40) | 7.99e-06 |
| (Missing) | 90 | 36 | 26 | 28 |  |
| t-Tau | 0.56 (0.60) | 0.53 (0.55) | 0.54 (0.57) | 0.77 (0.88) | 0.198 |
| (Missing) | 89 | 35 | 26 | 28 |  |
| GFAP | 5.32 (0.50) | 5.26 (0.49) | 5.46 (0.44) | 5.67 (0.47) | 4.57e-04 |
| (Missing) | 89 | 35 | 26 | 28 |  |
| NfL | 3.09 (0.55) | 3.03 (0.51) | 3.36 (0.61) | 3.43 (0.65) | 5.69e-04 |
| (Missing) | 89 | 35 | 26 | 28 |  |
| Aβ<sub>40</sub> | 5.04 (0.21) | 5.04 (0.22) | 5.06 (0.18) | 5.05 (0.17) | 0.837 |
| (Missing) | 37 | 8 | 14 | 15 |  |
| Aβ<sub>42</sub> | 3.75 (0.21) | 3.76 (0.21) | 3.68 (0.18) | 3.73 (0.23) | 0.153 |
| (Missing) | 37 | 8 | 14 | 15 |  |
| **Cognitive domain** (Last available exam) |  |  |  |  |  |
| Language | 0.25 (0.65) | 0.45 (0.53) | -0.14 (0.56) | -0.30 (0.73) | 2.08e-17 |
| (Missing) | 9 | 3 | 2 | 4 |  |
| Memory | 0.31 (0.61) | 0.51 (0.46) | 0.04 (0.47) | -0.30 (0.77) | 4.02e-21 |
| Executive function | -0.01 (0.62) | 0.17 (0.54) | -0.34 (0.43) | -0.51 (0.70) | 4.16e-15 |
| (Missing) | 5 | 2 | 1 | 2 |  |
| MMSE | 27.61 (3.23) | 28.36 (1.73) | 27.29 (2.81) | 24.88 (5.76) | 5.84e-12 |
| (Missing) | 4 | 4 | 0 | 0 |  |
| **Dementia type code (n)** |  |  |  | 1=26, 2=3, 4=9,<br>5=1, 6=7, 7=2, 10=3 |  |

<sup>1</sup>Mean (SD); n (%)<sup>2</sup>One-way analysis of means; Pearson's Chi-squared test; Fisher's exact test. Dementia type code; 1= Alzheimer's disease (AD) without stroke, 2=AD with stroke, 4=mixed dementia (AD+vascular), 5=Frontotemporal dementia, 6= Dementia with Lewy bodies, 7= Dementia that does not fit any other Category (progressive), 10= Dementia – Uncertain. CN, cognitively normal; MCI, mild cognitive; eTIV, estimated total intracranial volume

### 3.2 Longitudinal brain atrophy trajectories

First, we identified 211 MRI features significantly associated with MCI and dementia (FDR<0.05) using a linear mixed effects model, adjusting for covariates (**sTable-2**). Based on these features(n=211) ALASCA analysis revealed three significant PCs, collectively accounting for approximately 95% of the total variance (PC1=75.81%, PC2=13.78%, and PC3=5.37%) (**Figure-2A**). PC1 demonstrated a clear pattern of progressive brain atrophy over time in individuals with MCI and dementia. PC2 exhibited a distinct trajectory: it was elevated in MCI patients compared to both CN and dementia groups, increased from the first to the second MRI scan, and then began to decline. PC3 appeared less interpretable, as the atrophy trajectories of MCI and dementia groups crossed over those of CN individuals, suggesting it may not reflect a meaningful clinical pattern.

**Figure 2.**
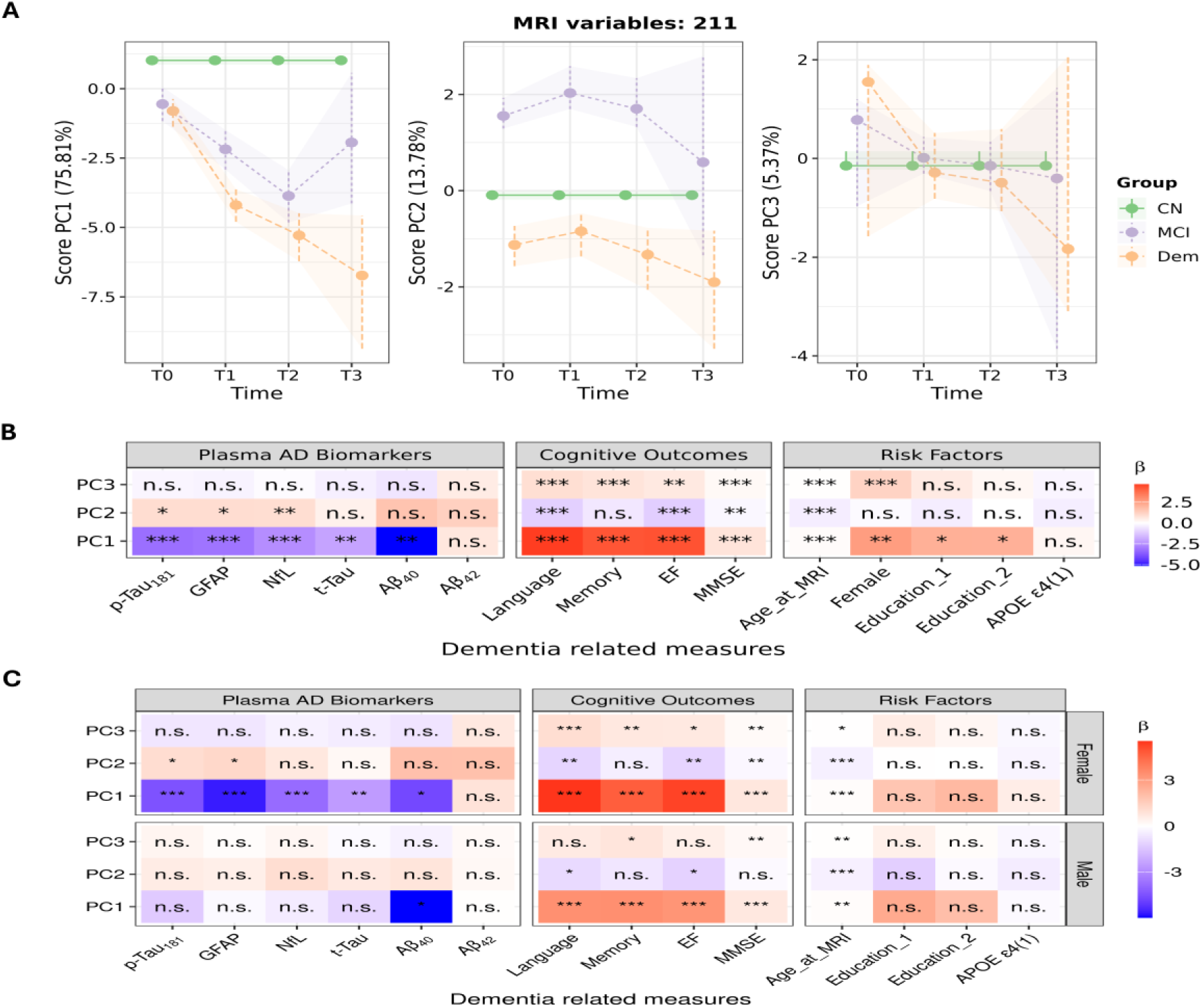
Principal components derived from the ALASCA group effect based on 211 significant MRI features. (**A**) Principal component (PC) scores representing the group effect across diagnostic categories. The CN group is shown as the reference, and differences in PC scores for MCI and dementia reflect disease-related structural deviations relative to CN. (**B**) Associations of PCs with plasma AD biomarkers and cognitive outcomes (**C**) Sex stratified sensitivity analysis. Association in Figure **B** and **C** were obtained using linear mixed-effects models of the form: MRI variable ∼ Biomarker (or cognitive score) + Age at MRI + sex + education + APOE ε4 + (1 | ID), where each MRI variable was treated as longitudinal outcomes and subject-specific random intercepts accounted for repeated measurements within individuals. Associations with demographic and genetic risk factors were evaluated using a multivariable model including all covariates simultaneously. Abbreviations: CN, cognitively normal; MCI, mild cognitive impairment; Dem, dementia; AD, Alzheimer’s disease; p-Tau_181_, phosphorylated tau 181; t-Tau, total tau; GFAP, glial fibrillary acidic protein; NfL, neurofilament light chain; Aβ_40_, amyloid-β40; Aβ_42_, amyloid-β42; EF, executive function, MMSE, Mini-Mental State Examination; MRI, magnetic resonance imaging; ALASCA, Assorted Linear functions for ANOVA Simultaneous Component Analysis. ***= 0<p-value<0.001, **=0.001< p-value <0.01, *=0.01< p-value <0.05, n.s=p.>0.05.

### 3.3 Characterization of the PCs relationships with plasma AD biomarkers and cognition

Among the plasma AD biomarkers, p-Tau181, GFAP, and NfL varied by diagnosis, while cognition strongly differed across CN, MCI, and dementia; other biomarkers were not significant(**Table-1**). PC1 is negatively associated with plasma AD biomarkers, including p-Tau_181_ (β=−3.21, p=1.66e-04), GFAP (β=−3.09, p=8.34e-05), NfL (β=−2.52, p=3.85e-04), and Aβ_40_ (β=−5.14, p=3.38e-03) (**sTable-4 and Figure-2B**). In contrast PC2 is modestly and positively associated with plasma AD biomarkers: p-Tau_181_ (β=0.91,p=2.71e-02), GFAP (β=0.90,p=1.85e-02), and NfL (β=0.89,p=1.00e-02). PC1 is positively associated with cognitive outcomes, such as language (β=4.64,p=1.50e-13), memory (β=4.22,p =1.85e-11), executive function (β=4.28,p=1.82e-11), and MMSE (β=0.73,p=8.02e-10) (**sTable S4 and Figure-2B**), whereas PC2 is negatively associated with cognitive outcomes, including language (β=−1.04, p=5.54e-02), executive function (β=−1.08,p=4.02e-04), and MMSE (β=−0.18,p=1.40e-03), but not with memory (**sTable-4 and Figure-2B**). PC1 was associated with age, sex, and education, whereas PC2 was associated with age only. Notably, PC1 showed a weak positive association with age (β=0.09, p=1.37e-06), while PC2 showed a strong negative association (β=−0.39, p=1.91e-106).

PC3 showed no significant associations with AD-related biomarkers and did not demonstrate a clear pattern across diagnostic groups. Although PC3 was modestly associated with age at MRI (β=0.04, p=6.55e-04) and sex (β=1.17, p=1.36e-05) (**sTable-4 and Figure-2B**), it did not exhibit consistent or biologically interpretable relationships with AD-related measures. Accordingly, we focused on PC1 and PC2 brain regions in our study. The loading values for 211 MRI variables for both PC1 and PC2 are provided in **sTable-3**. Results remained consistent in sex-stratified sensitivity analyses (**Figure 2C**), with generally stronger associations observed in females, consistent with prior literature.^33^

### 3.3 Characterization of important brain regions contributing to PC1 and PC2

The most robust brain regions for PC1 and PC2 were prioritized based on both high absolute loadings and narrow confidence intervals (CIs). For PC1, regions with loadings above the 60th percentile (|loading|>0.071) and CI widths below the 40th percentile (CI<0.032) were retained (**sTable-5**), identifying medial temporal structures most strongly associated with AD-related patterns. Key regions included hippocampal subfields (subiculum, CA1, molecular layer, dentate gyrus) and amygdalar nuclei (basal, lateral, accessory basal) (**Figure-3A**). For PC2, applying the same strategy (|loading|>0.064; CI<0.078) identified regions characterized by cortical gray–white matter contrast rather than volume (**sTable-6**). These were primarily distributed across frontal and temporal association cortices, including the frontal pole, orbitofrontal cortex, superior and middle frontal gyri, pars opercularis, supramarginal gyrus, and temporal pole (**Figure-3B**).

**Figure 3:**
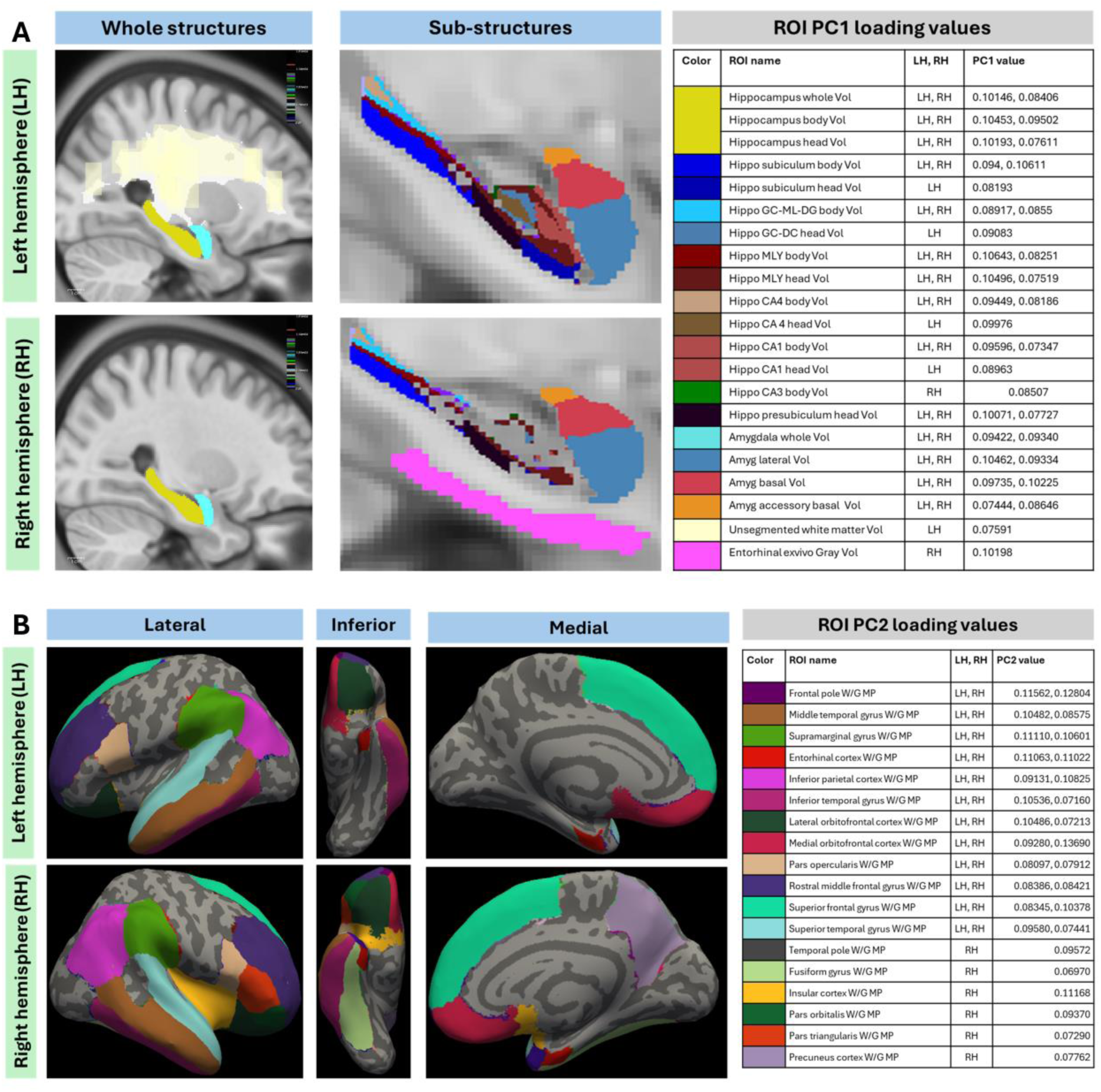
Most influential and robust brain regions contributing to PC1 (A) and PC2 (B). (**A**) Whole-structure and substructure regions of the hippocampus and amygdala contributing to PC1, selected using the percentile-based strategy described in the Methods. The adjacent table displays the corresponding PC1 loadings derived from the ALSCA model (see sTable-5). (**B**) Lateral, inferior, and medial views of white–gray matter contrast regions contributing to PC2, selected using the same percentile-based strategy. The adjacent table presents the corresponding PC2 loadings derived from the ALSCA model (see sTable-6). Volumetric regions identified for PC2 are not displayed here because they showed negative loadings (see sTable-6).

### 3.4 Association of PC1 and PC2 subregions with plasma AD biomarkers and cognitive outcomes

To further highlight the most biologically relevant regions, we identified the top five MRI features from each component based on the strength of their association with p-Tau_181_, the emerging biomarker of AD (**sFigure-3and4**). For PC1, the strongest associations were observed in the right entorhinal cortex (ex vivo gray matter volume), left subiculum head volume, and left molecular layer of the hippocampal head, as well as amygdalar nuclei such as the right basal nucleus and left lateral nucleus (**sFigure-2 and sTable-7**). Predicted longitudinal trajectory analyses revealed that these regions declined over time in individuals with MCI (slope range: −0.034 to −0.01) and showed a steeper decline in individuals with dementia (slope range: −0.069 to −0.018), whereas they remained relatively stable in CN individuals (slope range: −0.009 to 0.033) (**Figure-4A**). An exception was observed for the right entorhinal cortex, which showed a modest decline in individuals with dementia but an increasing trend in CN and MCI individuals (**Figure-4A**).

**Figure 4:**
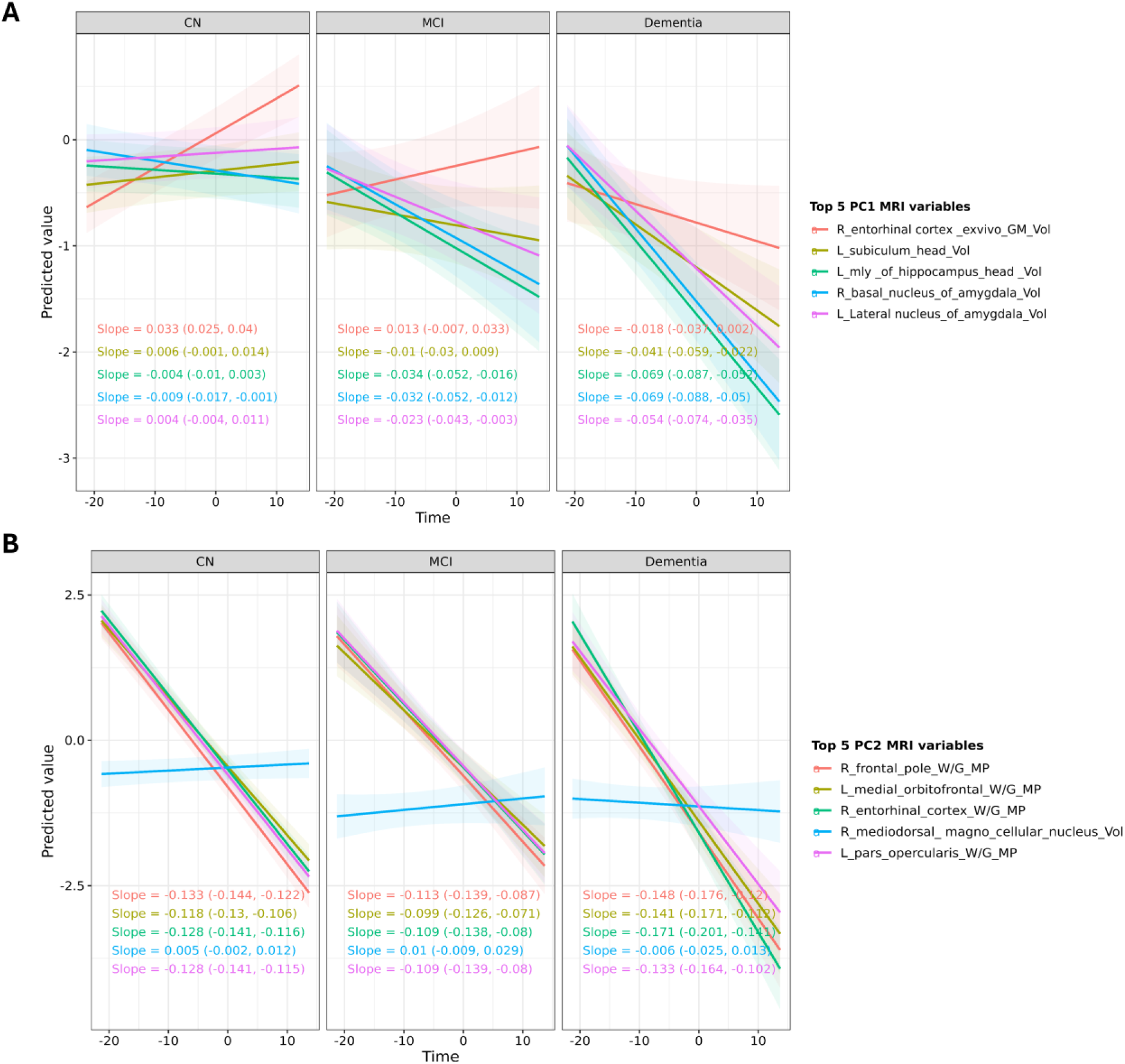
Predicted longitudinal trajectories of the top five PC1 (A) and PC2 (B) regions across diagnostic groups. Predicted trajectories for the top five MRI variables contributing to PC1 and PC2 were obtained from separate linear mixed-effects models including time since baseline MRI, diagnostic group, and their interaction, adjusted for sex, education, and APOE ε4 carrier status, with subject-specific random intercepts and random slopes for time. Time was defined as the interval from the first to the last MRI scan in CN individuals, and from the first MRI scan to disease onset in participants who developed MCI or dementia. Panels display model-based predicted values over time for each diagnostic group, with shaded regions representing 95% confidence intervals. Colored lines correspond to the top five PC1 and PC2 regions. Slopes (with 95% confidence intervals), derived from estimated marginal trends, are annotated within each panel to summarize the rate of change over time within diagnostic groups. Abbreviations: CN, cognitively normal; MCI, mild cognitive impairment.

The top PC2 regions selected using the same p-Tau_181_ based demonstrated a gradual and consistent decline across CN (slope range: −0.133 to −0.118), MCI (slope range: −0.113 to −0.099), and dementia (slope range: −0.171 to −0.133) groups, consistent with an overlapping aging and neurodegenerative process related structural pattern (**Figure-4B**). Whereas the right mediodorsal magnocellular nucleus volume remained relatively stable across diagnostic groups (slope: 0.005 in CN, 0.01 in MCI, and −0.006 in dementia). This region was therefore an exception and exhibited negative loading to PC2 (**Figure-4B and sTable-6**). Consistent with the predicted trajectories, inspection of the raw longitudinal trajectories for both PC1 and PC2 related regions observed similar pattern (**sFigure-4**).

### 3.5 Validation of PC1 top regions using brain neuropathology

Since PC1 captures brain regions most strongly associated with AD-related pathology, we used a subset of participants who donated brain tissue for neuropathological evaluation to assess whether top PC1 MRI regions were associated with Braak stage (neurofibrillary tangle pathology). This subset included 36 individuals with clinical diagnoses at the time of life (CN = 19, MCI = 6, dementia = 11), contributing a total of 104 longitudinal MRI scans (CN = 59, MCI = 17, dementia = 28)(**sTable-9**). Longitudinal trajectories of the top five PC1 MRI features were modeled as a function of Braak stage and time. The steepest volume loss occurred in individuals with advanced pathology (Braak 4–6; slope range:−0.097 to −0.011), whereas slow or no decline was seen in moderate and mild stages (Braak 2–3; slope range:−0.045 to 0.032; Braak 0–1; slope range:−0.027 to 0.037) (**Figure-5A**), supporting neuropathological validation of PC1 as an AD-related trajectory.

**Figure 5:**
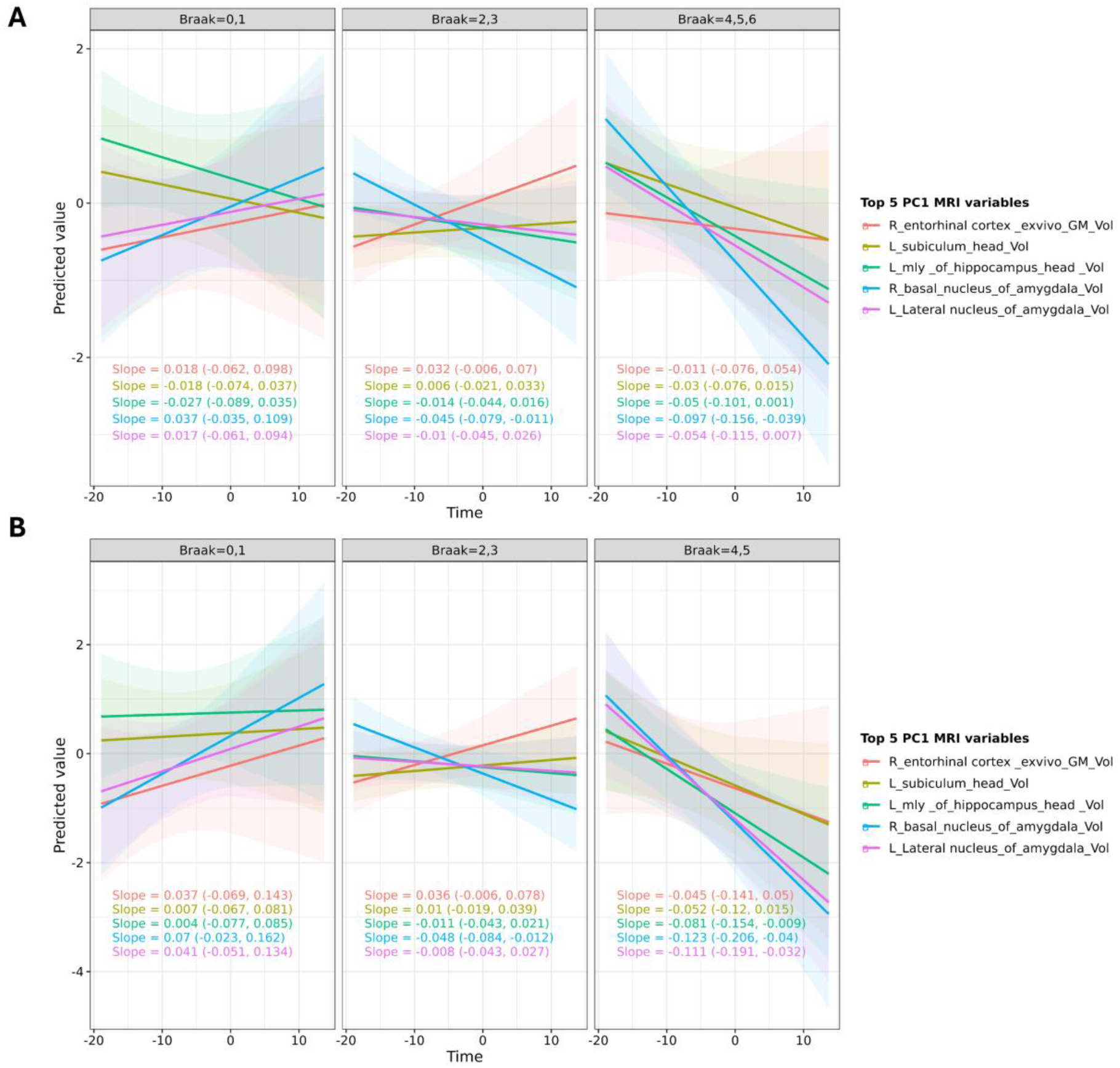
Predicted longitudinal trajectories of the top five PC1 regions with LATE positive individuals (A) and without LATE positive individuals (B) across different Braak stagings. Predicted trajectories for the top five MRI variables contributing to PC1 were obtained from separate linear mixed-effects models including time since baseline MRI, Braak staging group, and their interaction. Time was defined as the interval from the first to the last MRI scan in CN individuals, and from the first MRI scan to disease onset in participants who developed MCI or dementia. Panels display model-based predicted values over time for each Braak staging group, with shaded regions representing 95% confidence intervals. Colored lines correspond to the top five PC1 regions. Slopes (with 95% confidence intervals), derived from estimated marginal trends, are annotated within each panel to summarize the rate of change over time within Braak staging group. Abbreviations: CN, cognitively normal; MCI, mild cognitive impairment.

To evaluate robustness, we excluded seven individuals with co-existing LATE-NC pathology (stage1-3). After exclusion, the Braak 0–1 group no longer showed decline (slope range:0.004 to 0.041), the Braak 2-3 group demonstrated similar rates (slope range:−0.048 to 0.036), and the Braak 4–5 group exhibited steeper decline (−0.123 to −0.045) (**Figure-5B**). Notably, right entorhinal cortex atrophy was mild in the presence of LATE-NC (slope=−0.011; CI=−0.076 to 0.054) but became more pronounced after exclusion (slope=−0.045; CI=−0.141 to 0.05). No Braak 6 cases remained after removing LATE-NC, suggesting that cases with mixed LATE-NC accelerates the AD brain pathology to a severe level. Collectively, these findings indicate that the top five PC1 regions are more specific to pure AD (Braak) pathology.

## 4 Discussion

In this longitudinal, community-based study, we identified distinct trajectories of brain structural change using a data-driven framework (ALASCA). Two principal components captured biologically and clinically meaningful patterns. PC1 reflected progressive medial temporal neurodegeneration and showed strong associations with plasma AD biomarkers, cognitive decline, and Braak stage (**Figures 2-5**), whereas PC2 captured diffuse cortical gray–white matter contrast alterations reflecting more distributed structural changes (**Figures 2-4**). Together, these findings identify biologically anchored neurodegeneration trajectories and highlight the value of integrating MRI-derived change with blood-based AD biomarkers, cognition, and neuropathology.

### PC1: A longitudinal medial temporal–hippocampl/amygdalar neurodegeneration trajectory

PC1 represents a coherent trajectory of medial temporal neurodegeneration consistent with AD-related pathology. It accounted for more than 75% of the variance in longitudinal MRI change (**Figure-2A**) and was strongly associated with plasma p-Tau_181_, GFAP, and NfL(**Figure-2B**). These findings support the concept that AD-related neurodegeneration can be captured as a unified longitudinal pattern rather than isolated regional measures^34–36^ Prominent contributions from hippocampal subfields including the subiculum, CA1, molecular layer, and dentate gyrus align with known early vulnerability to tau pathology during Braak stages I–III.^37–39^ Consistent with prior studies that subregional medial temporal measures outperform whole-structure volumes in tracking AD progression^13,40^, PC1 brain regions underscore the value of subfield-resolved longitudinal MRI in clinical diagnosis and prognosis of AD. Neuropathological analyses further support the biological relevance of PC1. Exclusion of individuals with concomitant LATE-NC was associated with greater medial temporal decline, particularly the entorhinal cortex (**Figure-5A and 5B**), suggesting tighter coupling between medial temporal atrophy and AD pathology in the absence of mixed disease. Several individuals with mixed AD and LATE-NC pathology were younger at death and showed relatively preserved brain volumes, indicating heterogeneity in disease trajectories and possible survival-related effects(**sFigure-5**). Consistent with prior reports, LATE-NC commonly co-occurs with AD and contributes to variability in hippocampal and amygdalar atrophy.^41,42^

In addition to hippocampal involvement, PC1 prominently implicated amygdalar nuclei within the basolateral complex, including basal, lateral, and accessory basal regions. These nuclei are closely interconnected with hippocampal and entorhinal structures and are affected early by tau pathology and play key roles in memory and emotional processing.^14,37,43–45^ Prior imaging studies indicate that nuclei-specific amygdalar measures improve discrimination and differential diagnosis of AD^14,45,46^, and are associated with cognitive and neuropsychiatric severity, with the accessory basal nucleus showing particular vulnerability.^14,41^

### PC2: Diffuse cortical gray–white matter contrast alterations

In contrast to PC1, PC2 reflects diffuse cortical gray–white matter contrast alterations that do not align with classical medial temporal AD atrophy patterns. Instead, it captures distributed changes across frontal and temporal cortices, including the frontal pole, orbitofrontal cortex, and superior and middle frontal gyri (**Figure-4B**). Reduced gray–white matter contrast has been linked to microstructural processes such as myelin degradation, microvascular pathology, and altered tissue composition, and may precede overt cortical thinning.^26,47,48^ PC2 captures distributed cortical changes likely influenced by overlapping aging-related and AD/ADRD-related processes.^26^ The predominance of association cortices, along with stronger associations with executive dysfunction and weaker associations with memory performance, suggests that PC2 reflects cortical vulnerability shaped by overlapping aging- and disease-related processes rather than a single underlying disease mechanism for AD. This interpretation is supported by evidence that gray–white matter contrast provides information complementary to volumetric measures and may be sensitive to early neurodegenerative change.^48–50^

### Limitations

Several limitations should be considered. Longitudinal MRI data were acquired over multiple decades, during which imaging protocols evolved, potentially introducing residual variability. Plasma biomarkers were measured at different exam cycles, leading to possible temporal misalignment with MRI. Neuropathological data were available only in a subset of participants. In addition, participants entered the study at different ages and contributed varying numbers of scans over unequal follow-up intervals. Although mixed-effects models account for unbalanced data, estimated trajectories may reflect a combination of within-subject change and between-subject differences. Additionally, inclusion of multiple correlated measures per region (e.g., thickness, volume, surface area, and contrast) as inputs in ALASCA may introduce redundancy and potential multicollinearity, potentially overweighting specific regions in the derived components. On the other hand, different measurements for the same region also validated the pathological impact from different aspects. Finally, the FHS cohort is predominantly of European ancestry, which may limit generalizability.

## 5 Conclusions

This study demonstrates that longitudinal, data-driven modeling of MRI trajectories can characterize distinct patterns of brain structural change associated with neurodegeneration. A major strength of this study is the integration of longitudinal MRI trajectories with plasma biomarkers, cognition, and neuropathology. PC1-related regions showed consistent associations with biomarkers and cognitive performance, supporting their role as markers of disease severity.^51,52^ In contrast, PC2-related regions support that AD brains are biologically older than their chronological brain age. Accumulating evidence suggests that AD is associated with accelerated brain aging, in which neurodegenerative changes reflect amplification of biological processes that also occur during normative aging.^53,54^ Together, these components suggest that AD involves both focal medial temporal neurodegeneration (PC1), reflected by associations with amyloid- and tau-related biomarkers, and more diffuse, aging-related cortical vulnerability (PC2), consistent with processes such as neuroinflammation, metabolic dysfunction, and potential vascular contributions in the white matter of the brain.^55^ By integrating structural MRI with plasma AD biomarkers, cognitive outcomes, and postmortem neuropathology, this multimodal, trajectory-based framework may improve disease characterization, staging, and risk stratification in the clinical trials of AD.

## Acknowledgement

We want to express our thanks to all FHS participants for their decades of dedication and to the FHS staff for their hard work in collecting and preparing the data. We want to thank Dr. Phoebe Scollard for her insightful comments on this manuscript.

## Conflict of interest

The authors declare no conflict of interest. Rhoda Au is a scientific advisor to Signant Health and NovoNordisk.

## Funding

This study was supported by the Framingham Heart Study’s National Heart, Lung, and Blood Institute contract N01-HC-25195; National Institute on Aging grants U19-AG068753, RF1AG075832-01A1, U01-AG072577, R01-AG080810, and Framingham Heart Study Brain Aging Program (FHS-BAP) pilot grant by U19-AG068753; National Science Foundation grant DMS/NIGMS-2347698.

## Consent Statement

Informed consent was obtained from all study participants, and the Institutional Review Board of Boston University approved the study protocol.

## Data availability

Data set used in the preparation of this manuscript provided by the FHS-BAP and data is available on request. Please visit FHS-BAP website for more information https://www.bumc.bu.edu/fhs-bap/

## Notes

### Funding Statement

This study was supported by the Framingham Heart Study, National Heart, Lung, and Blood Institute contract N01-HC-25195; National Institute on Aging grants U19-AG068753, RF1AG075832-01A1, U01-AG072577, R01-AG080810, and Framingham Heart Study Brain Aging Program (FHS-BAP) pilot grant by U19-AG068753, National Science Foundation grant DMS/NIGMS-2347698.

