## Supplement Methods and Figure for "Longitudinal Brain Atrophy Patterns in Dementia and Cognitive Decline: the Framingham Heart Study"

[11]Athinoula A. Martinos Center for Biomedical Imaging, Massachusetts General Hospital, Boston, MA, USA
[12]Department of Psychiatry, Boston University Chobanian & Avedisian School of Medicine, Boston, MA, USA
[13]VA Boston Healthcare System, Boston, MA, USA
[14]Department of Medicine, University of Washington, Seattle, WA USA
[15]Department of Radiology and Imaging Services, Indiana University, Indianapolis, IN USA
[16]Indiana Alzheimer's Disease Research Center, Indiana University, Indianapolis, IN USA
[17]Department of Psychiatry and Behavioral Sciences, University of Washington, Seattle, WA USA
[18]VA Puget Sound, Geriatric Research Education and Clinical Center
*Co-corresponding authors
Wei Qiao Qiu,
Xiaoling Zhang,
Douglas N. Greve,

**sMethod**

**S1. Study participants**

Data for this study were obtained from participants in Generation 1 (Original) and Generation 2 (Offspring) cohorts of the Framingham Heart Study (FHS), a community-based, prospective cohort study of health in Framingham, Massachusetts. Participants with available structural longitudinal MRI data (n=1,256) processed using FreeSurfer-V 7.5 software at Massachusetts General Hospital were included. All participants underwent standardized cognitive assessments and were followed longitudinally until October 2024 (**Figure-1**). Detailed cohort design and selection criteria have been described previously.[1]

A total of 600 samples were excluded based on the following criteria: participants with missing APOE genotype (n=52), and missing diagnostic status (n=4), only one MRI scan (n=377), no individual with incident dementia had more than four scans (n=15; only 12 cognitively normal (CN) and 3 mild cognitive impairment (MCI) remains with more than 4 scans), and CN participants younger than 50 years (n=15). After exclusions, the final sample included 726 MRI scans from 300 participants (**Figure-1; sTable-1**). Each participant contributed at least two MRI scans (mean 2.52 scans per participant; range 2–4), reflecting repeated longitudinal assessments (**sTable-1**). Because inclusion required repeated MRI measures and complete clinical characterization, the analytic sample may be enriched for individuals with longer follow-up, older age, and greater data completeness. As a result, the study population may not fully reflect the broader FHS cohort, and longitudinal patterns may be influenced by selection and survivorship effects. Informed consent was obtained from all participants, and the Institutional Review Board of Boston University approved the study protocol.

**S2. Cognitive assessment and diagnostic adjudication**

Surveillance for cognitive impairment and incident dementia in the FHS began in 1976–1979 during the second half of their 14$^{th}$ health examination and the first half of their 15$^{th}$ health examination for the Original cohort and in 1979 for the Offspring cohort, beginning at the second health examination when participants were relatively young (mean [range] age 44 [17–77] years).[2,3] Since the mid-1980s, diagnoses of dementia and its subtypes have been determined through consensus adjudication by a panel including at least one neurologist and one neuropsychologist.

Cognitive screening and evaluation evolved over time. The Mini-Mental State Examination (MMSE) was administered as part of routine health examinations (beginning in 1981 for the Original cohort and 1991 for the Offspring cohort) and served as a screening tool to detect cognitive decline.[4] The Montreal Cognitive Assessment (MoCA) was administered in a limited number of examination cycles and was not part of the core longitudinal neuropsychological testing protocol. In contrast, comprehensive neuropsychological (NP) testing was conducted through ancillary examinations, initially targeting participants flagged for possible cognitive impairment and later expanded (beginning in 1999) to broader cohort participation.[5]

Participants meeting screening thresholds were referred for detailed clinical evaluation. The dementia review panel determined cognitive status, dementia subtype, and estimated date of onset using longitudinal neurologic examinations, detailed neuropsychological testing, medical records, and structured interviews with participants and informants.[6] Neuroimaging data were not used in diagnostic adjudication to avoid circularity with imaging-based analyses. Dementia was diagnosed according to DSM-IV criteria, and AD was classified based on NINCDS-ADRDA criteria. MCI was defined as cognitive decline not meeting criteria for dementia and was further classified by subtype and affected cognitive domains.[7,8]

Domain-specific cognitive factor scores for memory, language, and executive function were derived from neuropsychological test data using psychometric methods described previously, including confirmatory factor analysis and co-calibration across studies and test batteries to place scores on a common metric.[9] This approach accounts for variation in test availability and improves comparability across examination cycles. Scores with high measurement uncertainty (standard error >0.6) or derived solely from MMSE were excluded to minimize ceiling effects and measurement instability.[10] For association analyses with longitudinal MRI-derived measures, the most recent available cognitive factor scores and MMSE score were used. This approach provides a stable summary of cognitive performance and approximates cognitive status closest to the time of MCI or dementia onset, while avoiding complexities introduced by irregular cognitive testing intervals.

**S3. MRI Processing and feature extraction**

For the FHS, structural T1-weighted MRI scans were processed using the FreeSurfer – V 7.5 image analysis suite (https://surfer.nmr.mgh.harvard.edu). Participant confidentiality was ensured using the MiDeFace defacing procedure, which replaces identifiable facial features with an average face template ("MIDEFACE" label) while minimally altering non-facial neuroanatomy. As part of the centralized quality assurance (QA) pipeline, all scans were manually inspected to verify the presence of the MIDEFACE label and characteristic facial ridges, indicating that reconstructed images were non-identifiable. Overall image quality was assessed using a standardized 10-point visual rating scale (info_overall_score); scans scoring ≥6 were retained, while scans scoring ≤5 were excluded due to inadequate image quality or excessive correction requirements. Automated segmentation yielded volumes of whole-brain tissue, ventricles, cerebellum, brainstem, and subcortical structures, including the hippocampus and amygdala.[11]

Cortical thickness, surface area, and regional gray matter volume were extracted using the Desikan–Killiany and Destrieux atlases.[12,13] Hippocampal subfields, amygdalar nuclei, thalamic nuclei, and limbic structures were segmented using validated atlas-based, Bayesian, and deep learning approaches.[14-17] Each participant's MRI data then underwent intensity normalization, skull stripping, and automated segmentation of cerebral white matter to identify the gray matter/white matter boundary using FreeSurfer. Defects in cortical surface topology were automatically corrected, and the gray–white matter boundary was deformed outward to generate an explicit representation of the pial surface. All cortical surface reconstructions were visually inspected for technical accuracy and manually edited when necessary. Cortical thickness was calculated as the closest distance between the gray–white matter boundary and the gray–CSF boundary at each vertex on the tessellated cortical surface.

Gray–white matter tissue contrast was quantified using FreeSurfer's pctsurfcon output, which provides percent white/gray contrast values for cortical ROIs defined by the Desikan–Killiany aparc parcellation.[18] White–gray contrast was computed at each vertex on the cortical surface as $100 \times (W - G) / [(W + G) / 2]$,where W represents white matter signal intensity sampled 1 mm into the white matter from the white surface, and G represents gray matter signal intensity sampled at the midpoint of the cortical ribbon.[19] Vertex-wise contrast values were then averaged within each cortical ROI to generate region-level percent white–gray contrast measures. These sampling locations were selected to minimize partial volume effects while avoiding boundary crossings between tissue classes.

**S4. Plasma AD biomarker measurement**

Plasma biomarkers were measured at Offspring exams 07–09. Biomarkers included p-tau$_{181}$ (exam 09), t-tau, GFAP, and NfL (exam 08), and Aβ$_{40}$ and Aβ$_{42}$ (exam 07). Samples were analyzed using the Quanterix Simoa 2.0 platform.[3,20] Samples were stored at −80°C and thawed for 30–60 minutes at room temperature prior to analysis. Gentle inversion ensured homogeneity. Biomarkers were quantified at femtomolar concentrations. Log transformation and outlier evaluation were performed prior to analysis.

**Instrument and software**

- Simoa HD-1 Analyzer (Instrument ID: 27100100020; software version of 1.5)
- All samples was tested on the same instrument during the study.

**Sample handling**

- The mixed samples were quickly spun for 30s using a bench-top mini centrifuge to remove any liquid from the caps. A proper volume of the samples (100 µL of samples run in singlet; 130 µL of samples run in duplicate) were then transferred to 1.7-mL micro-centrifuge tubes pre-labeled with barcodes that match the original sample tubes respectively.
- The transferred samples were then centrifuged at 20,000g for 3 minutes at 4C.
- After centrifugation, the samples were transferred to 96-well plates following a pre-defined plate map for testing. All samples were diluted 4-fold automatically on the HD-1 Analyzer using sample diluent (based on recommended dilution scheme in kit package insert).
- Original samples were immediately refrozen after the testing aliquot was removed.

**Preparation of calibrators and controls**

- Ready-to-use calibrators were manufactured and frozen (-80C) at the following Tau concentrations: 0, 0.093, 0.298, 1.04, 3.00, 9.38, 30.5, 95.1 pg/mL.
- Ready-to-use controls with assigned ranges were pre-manufactured and stored frozen at -80C.
- The same lot of calibrators and controls were used during this study.

- Controls were treated as samples and diluted 4-fold automatically on the HD-1 Analyzer.

**Assay characteristics**

| Assay characteristics | p-Tau | t-Tau | Aβ40 | Aβ42 |
|---|---|---|---|---|
| Number of measurements (single/duplicate) | 12% run in duplicate | 11.6% in duplicate | 9% in duplicate | 9% in duplicate |
| Intra-assay CV | 5.10% | 4.10% | 3.20% | 2.60% |
| Inter-assay CV | 9.60% | 7.50% | 10.50% | 7.60% |
| Phantom samples | ICC = 0.992; CV=7.9%; N=130 | ICC = 0.741; CV = 9.2%; N = 324 | ICC = 0.916; CV = 4.8%; N = 146 | ICC = 0.943; CV = 3.5%; N = 146 |

**S5. Neuropathology**

AD-related neuropathology was assessed in a subset of FHS participants who donated brain tissue for postmortem evaluation. Neurofibrillary tangle pathology was quantified using Braak staging, which classifies the topographical extent of tau pathology from transentorhinal to neocortical regions. Neuropathological evaluations were conducted by board-certified neuropathologists at the Boston University Alzheimer's Disease Research Center (BU-ADRC), who were blinded to all demographic and clinical information.[21,22] In addition, phosphorylated TDP-43 (p-TDP-43) pathology was assessed using immunohistochemistry on sections including the amygdala with entorhinal cortex, hippocampus, and dorsolateral frontal cortex. Limbic age-related TDP-43 encephalopathy (LATE) neuropathologic change was defined according to established consensus criteria.[23,24]

**S6. Statistical analyses**

All statistical analyses were conducted in R (version 4.4.0). Continuous variables were summarized as mean ± standard deviation (SD), and categorical variables were summarized as counts and percentages. Baseline differences across diagnostic groups (cognitively normal [CN], mild cognitive impairment [MCI], and dementia) were evaluated using one-way analysis of variance (ANOVA) for continuous variables and Pearson's chi-square tests for categorical variables. To control for multiple comparisons, false discovery rate (FDR) correction was applied, with FDR-adjusted $p < 0.05$ considered statistically significant.

**Feature selection using linear mixed-effects models**

To identify MRI-derived structural features associated with diagnostic status while accounting for repeated measurements, separate linear mixed-effects models were fitted for each MRI feature. Participant-specific random intercepts were included to account for within-subject correlation across repeated MRI scans, *MRI feature ~ diagnostic group (ref=CN, MCI, dementia) + Age at MRI + sex + education + APOE ε4 + (1 | ID)*. These models tested for overall differences in MRI measures across diagnostic groups while adjusting for relevant covariates age at MRI, sex, education, and APOE ε4 carrier status. MRI features meeting FDR $< 0.05$ were retained for subsequent trajectory modeling.

**ALASCA modeling framework**

Selected MRI features were entered into Assorted Linear functions for ANOVA–Simultaneous Component Analysis (ALASCA), implemented via the ALASCA R package.[25] ALASCA extends classical ANOVA–Simultaneous Component Analysis to longitudinal data by first modeling predefined experimental effects using linear mixed-effects models and then decomposing the resulting multivariate effect matrices into orthogonal principal components (PCs). The specified model was: *MRI value ~ Time * diagnostic group (ref=CN, MCI, dementia) + Age at MRI + (1 | ID)* where Time represented discrete time since baseline MRI and group corresponded to diagnostic category. The Time × group interaction enabled estimation of differential longitudinal trajectories across diagnostic groups. Features were scaled using the standard deviation at the first time point (scale_function = "sdt1") to standardize variance. Model robustness was assessed using jackknife validation (validate = TRUE) with 500 resampling iterations (n_validation_runs = 500, validation_method = "jack-knife"), from which confidence intervals for loadings were derived.

The number of retained PCs was determined based on proportion of explained variance and interpretability of the resulting longitudinal trajectories. Component scores were extracted for each participant and time point and used as summary measures of multivariate brain atrophy patterns.

**Association analyses**

Associations between ALASCA-derived PCs and plasma AD biomarkers and cognitive outcomes were evaluated using linear mixed-effects models:

*PC ~ Biomarker (or cognitive score) + Age at MRI + sex + education + APOE ε4 + (1 | ID)*

In these models, PC scores were treated as longitudinal outcomes, and predictors were treated as fixed variables. Random intercepts accounted for repeated PC measurements within individuals. Biomarker values were log-transformed when necessary to reduce skewness.

**Identification of influential regions**

To identify the most influential and robust regions underlying significant PCs, MRI features were evaluated based on both magnitude and stability of their ALASCA-derived loadings. The absolute loading value quantified the strength of regional contribution, while the width of the jackknife-derived confidence interval (CI) reflected robustness.

For anatomical interpretation of clinically relevant trajectories, associations between individual MRI features and plasma phosphorylated tau 181 (p-$tau_{181}$) were examined. For each PC, MRI features were ranked by strength of association with p-$tau_{181}$, and the top five features were selected for descriptive characterization. Longitudinal trajectories of these regions were visualized from the first to the last MRI scan in CN participants and from the first MRI to disease onset in individuals with MCI or dementia.

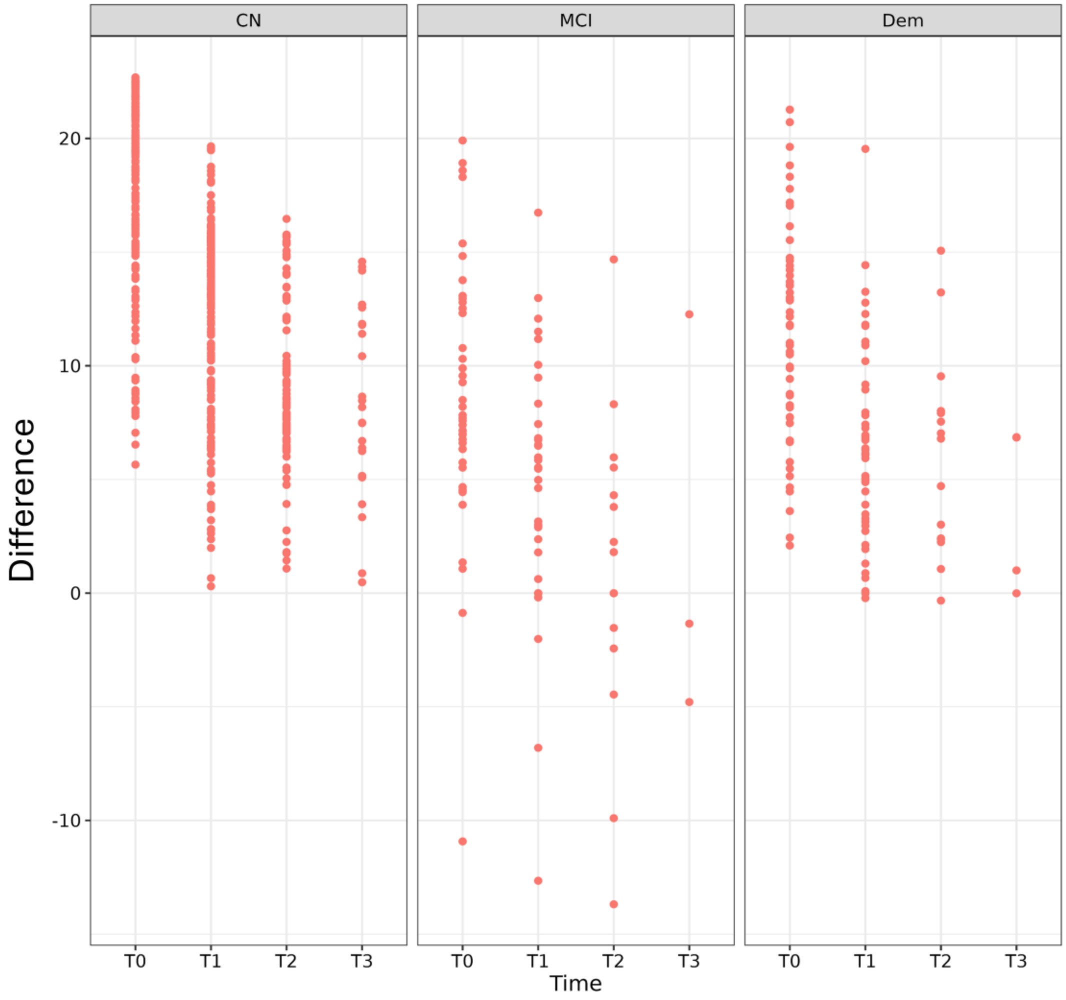


**sFigure-1**: Distribution of the difference between age at onset (for MCI and dementia) or age at last contact (CN participants) and age at MRI scan. Positive values indicate that the onset of MCI or dementia occurred after the MRI scan. All MCI and dementia cases represent incident diagnoses relative to the MRI scan, with only a few participants classified as MCI at the time of imaging. Abbreviations: CN, cognitively normal; MCI, mild cognitive impairment; Dem, dementia

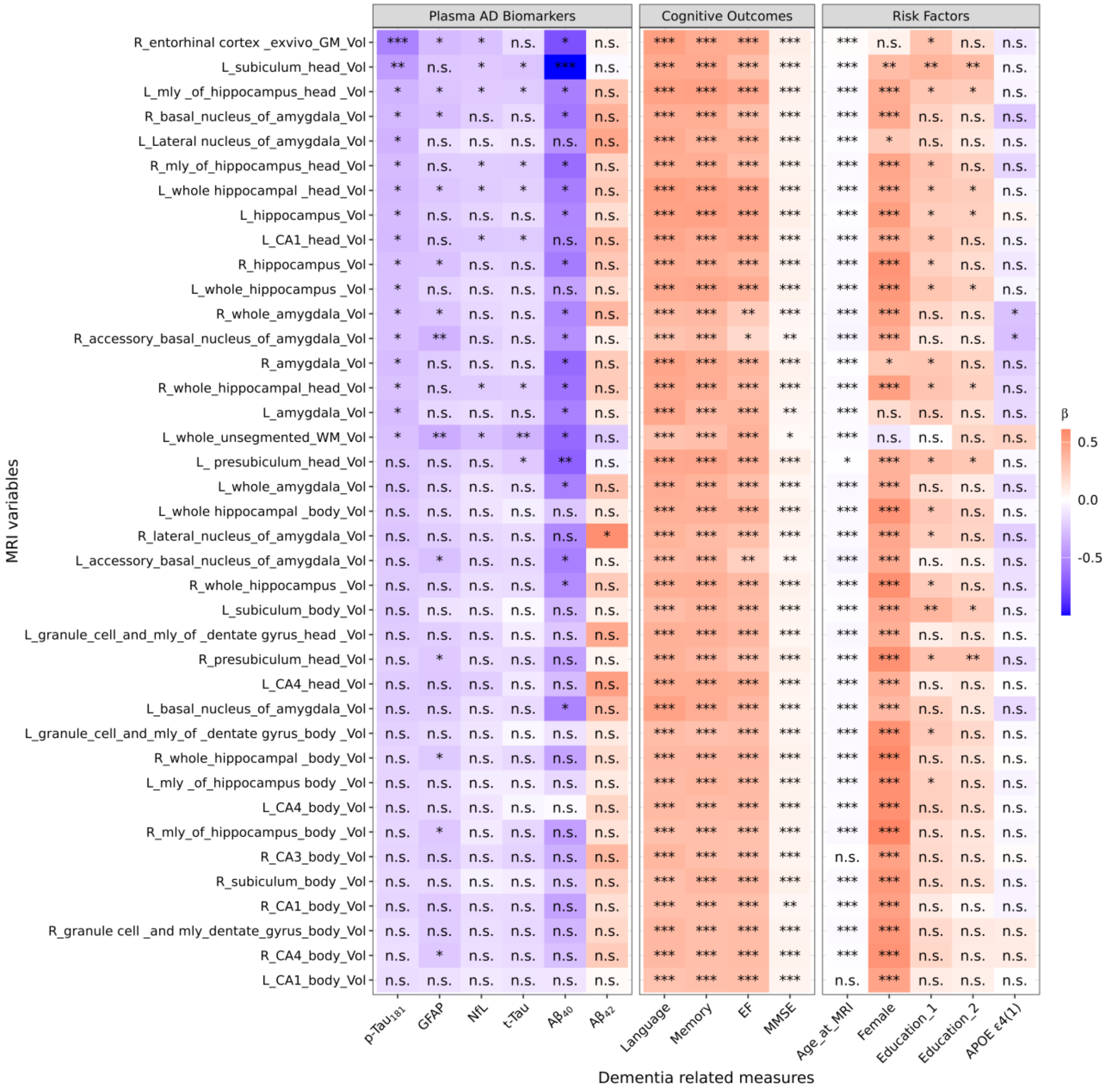


**sFigure-2**: Associations of **PC1** selected regions with plasma AD biomarkers and cognitive outcomes estimated using linear mixed-effects models of the form: *MRI variable ~ Biomarker (or cognitive score) + Age at MRI + sex + education + APOE ε4 + (1 | ID)*, where each MRI variable was treated as longitudinal outcomes and subject-specific random intercepts accounted for repeated measurements within individuals. Associations with demographic and genetic risk factors were evaluated using a multivariable model including all covariates simultaneously. This figure corresponds to Supplementary Table S7. Rows are ordered according to the p-Tau$_{181}$, from smallest to largest. Abbreviation: p-Tau$_{181}$, phosphorylated tau 181; t-Tau, total tau; GFAP, glial fibrillary acidic protein; NfL, neurofilament light chain; Aβ$_{40}$, amyloid-β$_{40}$; Aβ$_{42}$, amyloid-β$_{42}$; EF, executive function, MMSE, Mini-Mental State Examination. ***= 0<p-value<0.001, **=0.001< p-value <0.01, *=0.01< p-value <0.05, n.s.=p.>0.05.

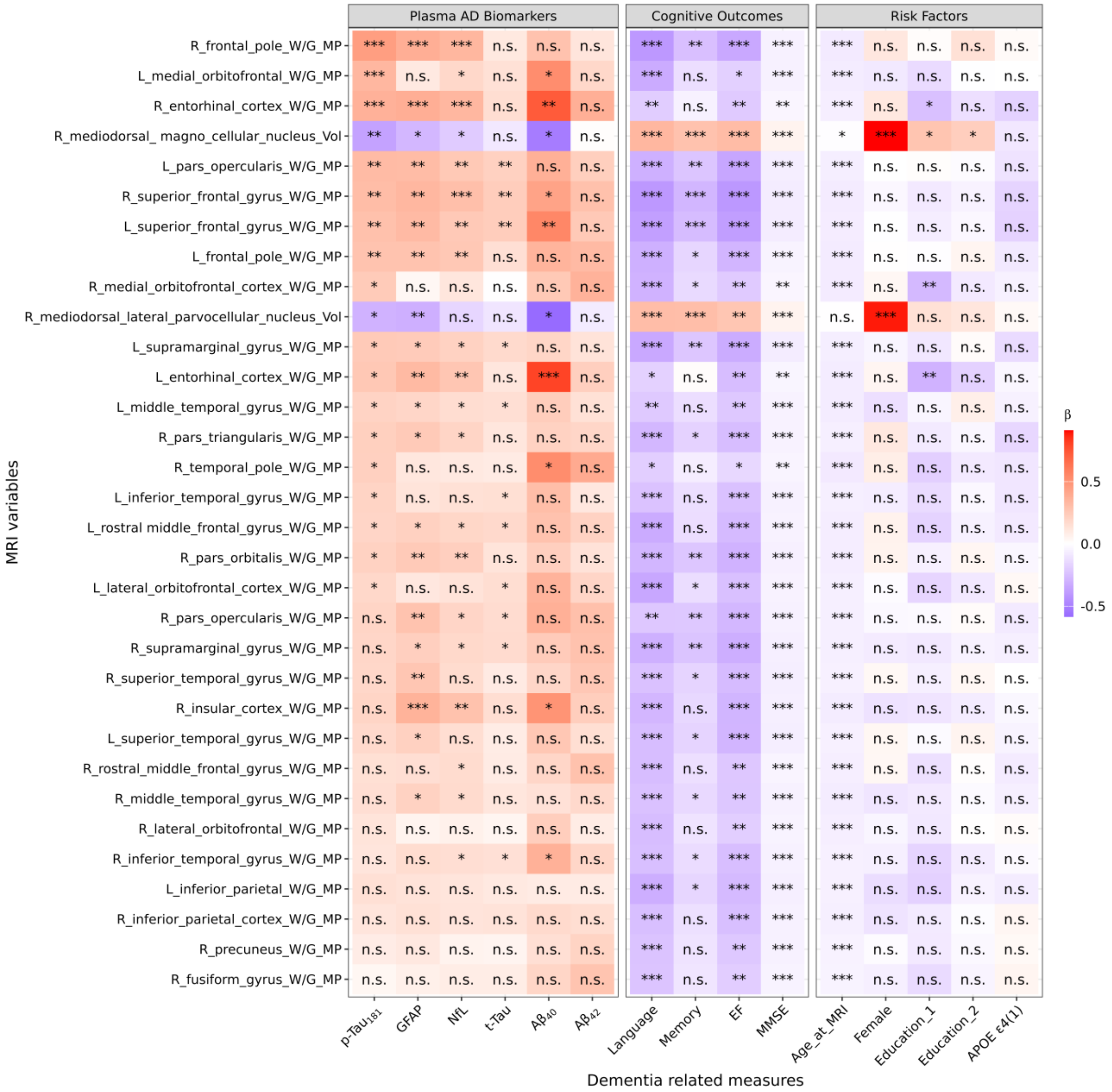


**sFigure-3**: Associations of **PC2** selected regions with plasma AD biomarkers and cognitive outcomes estimated using linear mixed-effects models of the form: MRI variable ~ Biomarker (or cognitive score) + Age at MRI + sex + education + APOE ε4 + (1 | ID), where each MRI variable was treated as longitudinal outcomes and subject-specific random intercepts accounted for repeated measurements within individuals. Associations with demographic and genetic risk factors were evaluated using a multivariable model including all covariates simultaneously. This figure corresponds to Supplementary Table S8. Rows are ordered according to the p-$Tau_{181}$, from smallest to largest. Abbreviation: p-$Tau_{181}$, phosphorylated tau 181; t-Tau, total tau; GFAP, glial fibrillary acidic protein; NfL, neurofilament light chain; $A\beta_{40}$, amyloid-$\beta_{40}$; $A\beta_{42}$, amyloid-$\beta_{42}$; EF, executive function, MMSE, Mini-Mental State Examination. ***= 0<p-value<0.001, **=0.001< p-value <0.01, *=0.01< p-value <0.05, n.s.=p.>0.05.

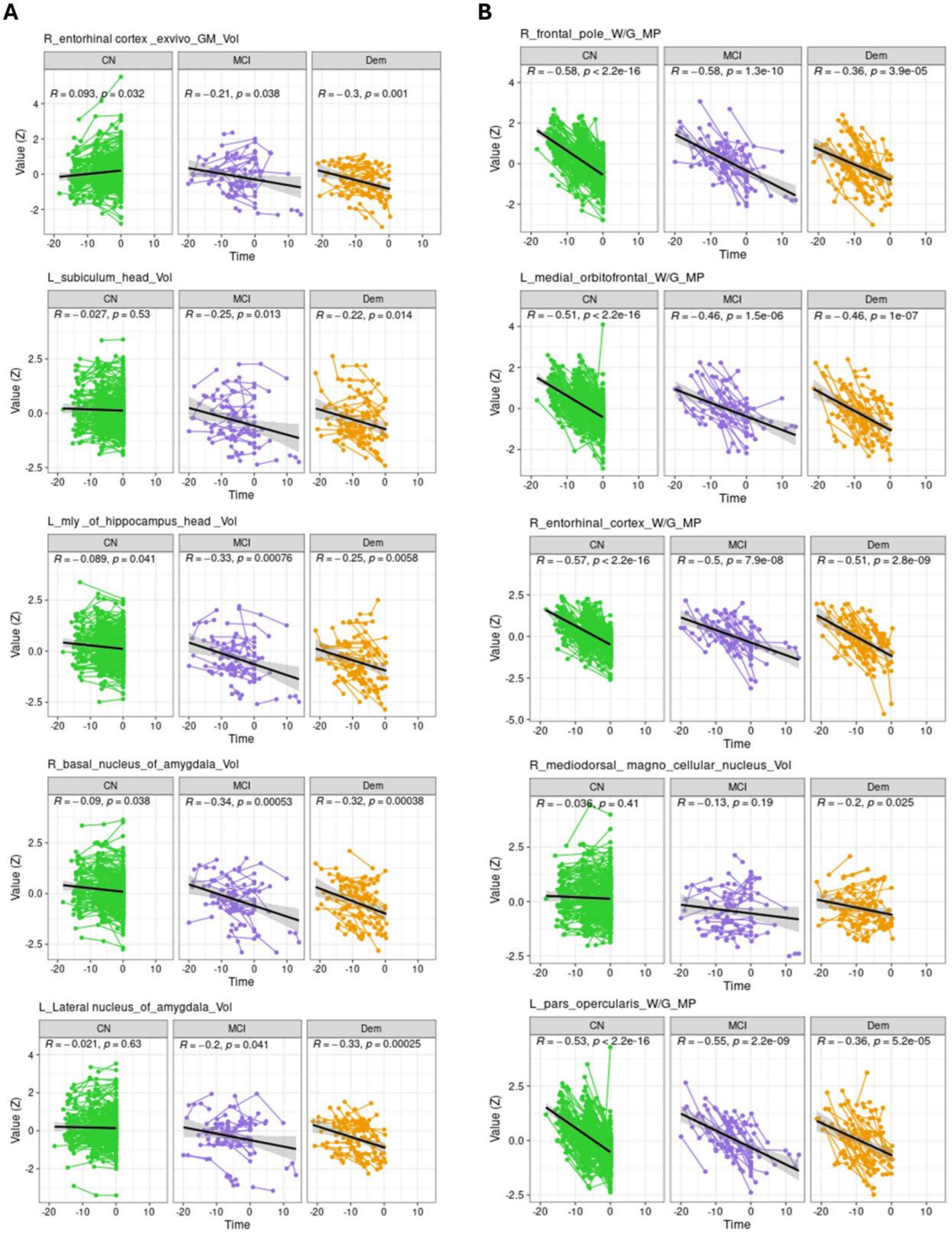


**sFigure-4**: Raw trajectories of top five PC1 (**A**) and PC2 (**B**) regions. Time was defined as the interval from the first to the last MRI scan in CN individuals, and from the first MRI scan to disease onset in participants who developed MCI or dementia

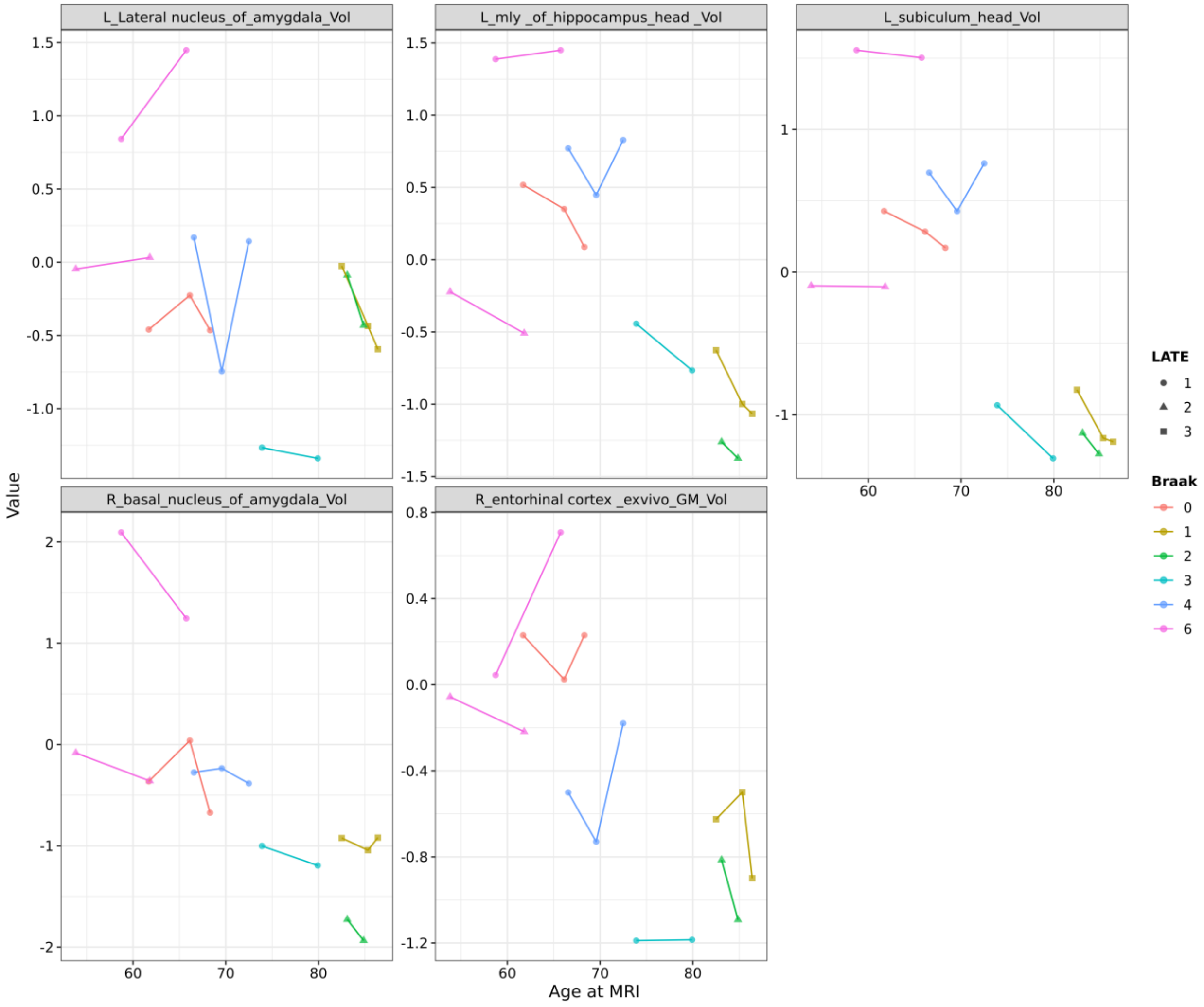


**sFigure-5**: Distribution of volumes of the five PC1 regions across age at MRI in seven individuals with LATE positivity.
